# Targeted reduction of airborne viral transmission risk in residential aged care

**DOI:** 10.1101/2022.05.30.22275789

**Authors:** Amanda Brass, Andrew P Shoubridge, Nicolas Larby, Levi Elms, Sarah K Sims, Erin Flynn, Caroline Miller, Maria Crotty, Lito E Papanicolas, Steve L Wesselingh, Lidia Morawska, Scott C Bell, Steven L Taylor, Geraint B Rogers

## Abstract

COVID-19 has demonstrated the devastating consequences of the rapid spread of an airborne virus in residential aged care. We report the use of CO_2_-based ventilation assessment to empirically identify potential “super-spreader” zones within an aged care facility, and determine the efficacy of rapidly implemented, inexpensive, risk reduction measures.

## Main Text

Outbreaks of COVID-19 in residential aged are facilities (RACF) have proven devastating. The combination of high transmission rates and resident vulnerability has led to rates of mortality that are higher than any other setting, including acute care hospital facilities [1]. In response, aged care providers and public health authorities have enacted measures to reduce the risk of SARS-CoV-2 exposure to prevent transmission within facilities. These measures include social distancing, reduced visitation by family members (site lockdowns), the wearing of personal protective equipment (PPE) by staff and visitors, and isolation of those with active infection. The impact of these measures on the well-being and quality-of-life of residents, more than half of whom have reduced cognitive ability or dementia [2], has been considerable [3]. The need to identify less disruptive measures to protect RACF populations from airborne viral transmission (including respiratory viruses other than SARS-CoV-2) is urgent.

Current strategies to reduce rates of indoor viral transmission can be placed into two broad categories; 1) those that reduce the susceptible occupant population, including occupancy limits, vaccination programs, and PPE use, and 2) those that prevent the accumulation of viable viral particles within the circulating air of an RACF environment [4]. This latter group of measures is more effective as it doesn’t require any behaviour changes to staff or visitors. It also offers long-term advantages by avoiding the need to place additional burden on facility residents and staff, or the need to limit access to facilities by family members. Several strategies to reduce the build-up of viable viral particles can be employed, including the use of high-efficiency particulate absorbing (HEPA) air-filtration, or ultraviolet C (UVC) disinfection devices [3,4]. However, most effective approach is arguably to remove viral particles through increased ventilation [5].

Outdoor venues are associated with a low relative risk for airborne viral transmission due to the rapid clearance of virus-containing airborne particles through natural air movement. Indoor spaces, where air exchange is considerably reduced, are estimated to have a SARS-CoV-2 transmission risk that is more than 18 times higher [6]. Increasing rates of air exchange within the built environment would reduce this transmission risk substantially. The principal reason that this strategy has not been employed widely is the considerable costs associated with heating or cooling external air [7]. Instead, many facilities, including RACFs, typically employ high rates of air recirculation, inadvertently resulting in recirculation of the virus as well [5].

Increasing ventilation within facilities that house vulnerable populations has been advocated widely during the COVID-19 pandemic [8]. However, implementation of ventilation measures has been limited. The ability to target increases in air exchange to those zones within facilities that carry the highest transmission risk would greatly increase the financial feasibility of improving ventilation. Here we report the use of a rapid, low-cost, strategy to empirically identify potential “super-spreader” zones within an RACF, and the efficacy of the targeted ventilation-based risk reduction measures that were deployed in response.

While atmospheric CO_2_ levels remain relatively constant, indoor CO_2_ levels increase as a result of human respiration. Elevated CO_2_ levels have been used widely as a marker of indoor air quality and, relative to occupancy, a measure of air exchange and ventilation efficiency. Indeed, such measures correlate strongly with infectious disease outcomes, such as absenteeism due to acute respiratory infection [9]. CO_2_ levels can be determined accurately using inexpensive monitors, allowing for high transmission risk zones within facilities to be identified rapidly and mitigation strategies to be enacted. We used 62 wall-mounted remotely-monitored CO_2_ sensors (Aranet4 Pro Sensors, Radical, Australia) to monitor CO_2_ levels over a four-week period within a two-storey 110-bed RACF in Australia. During this period, all staff and visitors to the RACF were required to wear N95 face masks (or equivalent), goggles or face shield, and be vaccinated against COVID-19. Deployment of CO_2_ monitors covered the majority of accessible sites within the facility, including “back of house” areas used by staff only (n=17) and mixed communal areas used by both staff and residents (n=45). The zones employed a range of heating, ventilation, and air-conditioning (HVAC) systems, including evaporative cooling with 100% fresh ducted air, refrigerated reverse cycle ducted air, refrigerated reverse cycle split system, and exhaust air. Each monitor also captured temperature, humidity, and pressure, enabling data normalisation.

Over the course of the survey period, none of the 45 mixed zones showed indications of poor ventilation, with the highest peak CO_2_ concentration being 830 ppm, which was in the resident dining room during the lunch period. However, of the 17 assessed back of house zones, which were only accessible to staff, 3 were identified as high transmission risk based on CO_2_ levels exceeding 1000 ppm for a period of 15 min or more (levels logged at 60 second intervals). These included one of the two staff lunch rooms and two of the three staff meeting rooms (**Figure 1A**). Interventions were identified based on the zone usage, the HVAC modality employed, and the ability to achieve improved ventilation rapidly and at low relative cost.

**Figure 1:**
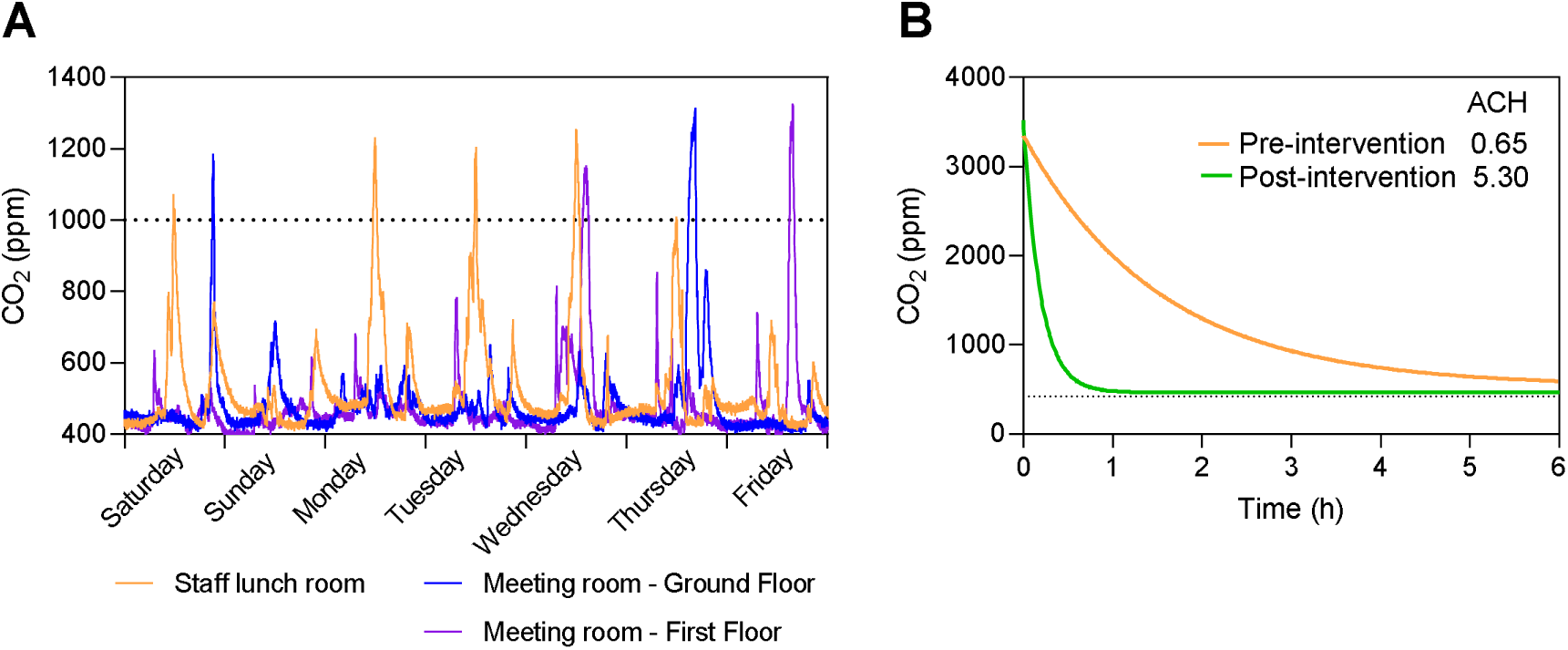
A) CO_2_ peaks over a one-week period for the identified high risk transmission zones. B) CO_2_ decay curve for the staff lunch room before (orange) and after (green) extraction fan installation. Air changes per hour (ACH) calculated by fitting non-linear regression.

### High risk zone 1

The staff lunch room had a floor area of 19 m^2^ with a refrigerated reverse cycle split system HVAC. Despite room density restrictions of 1 person per 4 m^2^, high, regular, and prolonged CO_2_ peaks were identified in the lunch room corresponding to meal breaks (**Supplementary Figure 1**). Of note, staff were not masked while eating and CO_2_ levels did not return to baseline levels for several hours after these peak periods. To quantify room ventilation, a subsequent CO_2_ decay curve was generated over a 6-hour period, during which the staff lunch room was not occupied. The resulting air changes per hour (ACH) was 0.65 (**Figure 1B**), which is 10-fold lower than the Australian Standards for ventilation in a healthcare setting [10]. Discussion with the facility’s environmental management team identified that installation of an extraction fan at a cost of $3400 AUD would effectively ventilate the room. The fan was set to extract air at 100 L/s and connected to a room sensor allowing usage to be limited to times of room occupancy. Retesting ventilation by CO_2_ decay curve after extraction fan installation yielded an improved ACH of 5.30 (**Figure 1B**). Ongoing running costs of the extraction fan were calculated by an AC current and voltage data logger to be $40.20 AUD per year.

### High risk zones 2 and 3

Two staff meeting rooms also had CO_2_ peaks indicative of high transmission risk. The first room had a floor area of 50 m^2^ and a refrigerated reverse cycle ducted air system controlled by a remote control. CO_2_ peaks in this room were less frequent than the staff lunch room, with a total of two peaks over 1000 ppm during the survey period. It was noted by facility management that the HVAC was rarely switched on during meetings, potentially explaining these CO_2_ peaks. Intervention strategies were therefore minimal, with the HVAC system programming adjusted to automatically turn on during times when the room is in use. The second room had a floor area of 20 m^2^ and a refrigerated reverse cycle split system HVAC. Like the first meeting room, a total of two peaks above 1000 ppm were recorded in this room during the survey period. However, unlike the first meeting room, existing HVAC could not be easily modified to improve ventilation. Interim protocols were therefore implemented, continuing a mandated mask use and room density limit for staff using this room.

Globally, the COVID-19 pandemic has again underlined the vulnerability of those living in residential aged care to airborne transmission of viral infections. In addition to COVID-19, seasonal respiratory viruses, including seasonal influenza and respiratory syncytial virus (RSV) represent a major cause of morbidity and mortality for residents [11]. Indeed, following a period of reduced exposure, the seasonal influenza peaks in 2022 are predicted to pose a significant health threat, as seen in the 2021-2022 winter in the Northern Hemisphere [12]. Targeting effective risk reduction measures to areas that are likely to contribute disproportionately viral spread could provide considerable protection.

In our analysis, passive and simulated CO_2_ levels were used to identify high transmission risk zones. Such approaches are simple and readily deployable, however do not account for other factors affecting transmission risk, such as PPE and level of interaction [13]. CO_2_ peaks were also based on levels recommended by The American Society of Heating, Refrigerating and Air-Conditioning Engineers (ASHRAE) of less than 600 ppm above outdoor concentrations (approximately, 420 ppm) [14]. The recommendation of these thresholds is based on consideration of a number of factors, including comfort. Alignment of target levels more closely with empirically assessed viral transmission rates would enable further protocol refinements.

The approach we describe has the potential to be applied beyond aged care. Rapid, targeted, improvements in air quality management, based on empirical facility surveys, could substantially reduce airborne pathogen transmission in other high-risk contexts, and warrant consideration in settings such as detention facilities, hospitals, or schools.

## Data Availability

All data produced in the present study are available upon reasonable request to the authors

---

**Supplementary Figure 1:**
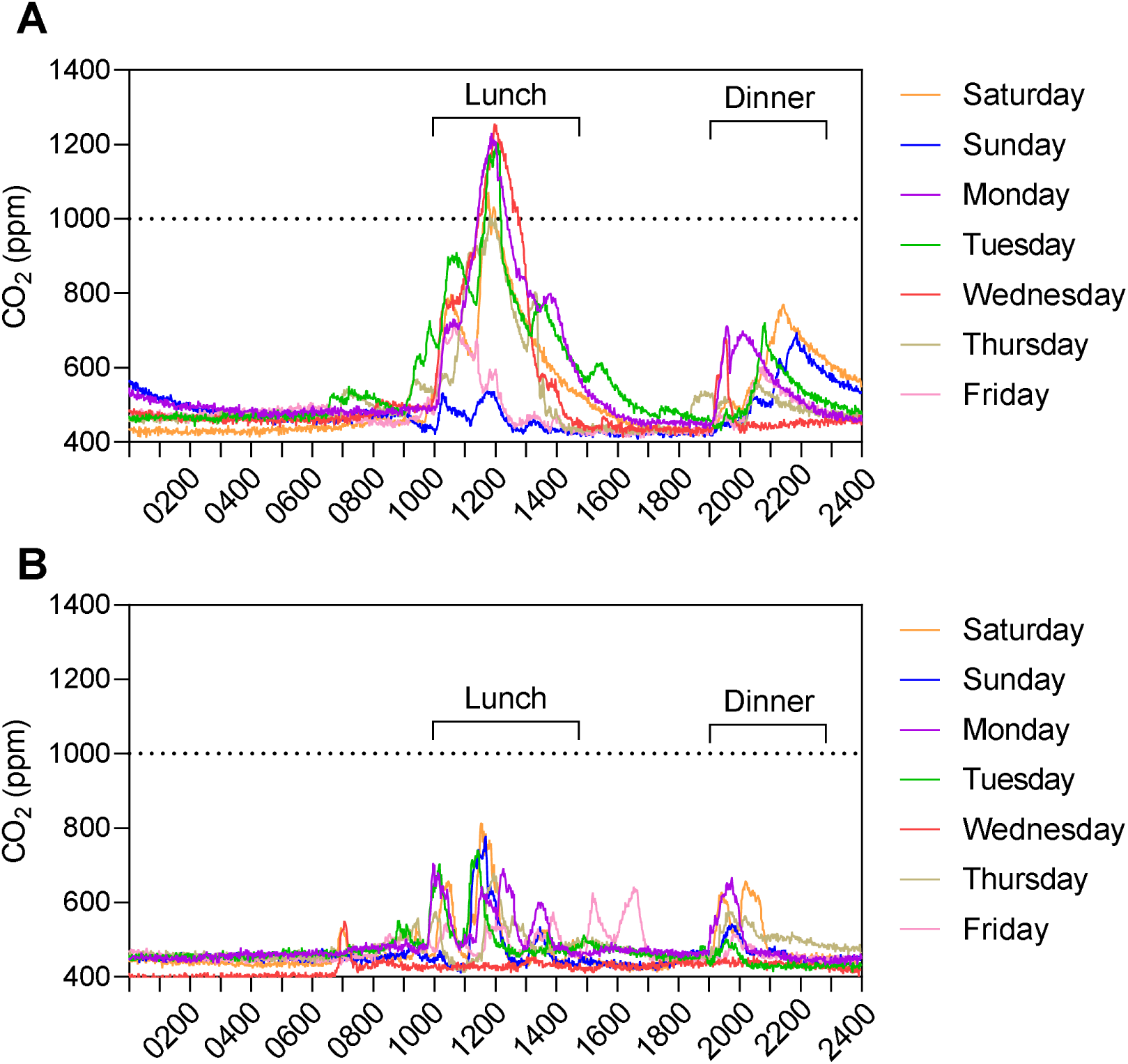
CO_2_ tracing of a 24 h period in the staff lunch room over the course of a week. A) Prior to the installation of the extraction fan, showing high prolonged peaks between 10am-2pm and 7pm-10pm, corresponding with staff lunch and dinner breaks. B) After the installation of the extraction fan, showing reduced peaks.

